# Irish Defence Forces graduate’s opinions and experiences of UCD’s Diploma in Military Medicine

**DOI:** 10.1101/2020.02.05.20020651

**Authors:** S. Loughman, C. Berry, Paul Hickey, Mark Ruddy, Brian Bruno, G. Bury

**Affiliations:** Centre for Emergency Medical Sciences, UCD, Dublin, Ireland; Former Head of Defence Forces Medical School, Curragh, Co. Kildare, Ireland; Defence Forces Medical Corps, Curragh, Co Kildare, Ireland; Advanced Paramedic Tutor, Centre for Emergency Medical Sciences, UCD, Dublin, Ireland

## Abstract

**Aims:** This study explores the opinions and experiences of Irish Defences Forces graduates from UCD’s Diploma in Military Medicine. It aims to identify which aspects of medical education are relevant for the development of military graduates in future.

**Methods:** A validated Clinical Learning Environment Score (CLES) tool was adapted and incorporated into an online survey. This was sent electronically to 71 graduates. Responses were anonymous.

**Results:** 38 (54%) of graduates responded. Just 16 (42%) use their new skills in their daily work. Of the 9 (24%) deployed overseas, all used their new skills. Emergency and Occupational Health skills were used more, while advanced skills were used rarely. Only 22 (40%) of DMMC graduates work in the Defence Forces Medical Unit.

**Conclusion:** Student feedback was positive regarding teaching and clinical placements at UCD. An emphasis on frequently used skills, and a change in how Defence Forces acknowledge qualifications, may support more graduates in entering the military medical workforce.

**Key Messages:**

- The Irish Defence Forces identified the need for an innovative training programme to equip its non-medical staff to address the role known as ‘Combat Medical Technician’(CMT).
- This led to the development of the UCD Diploma in Military Medical Care (DMMC) programme.
- The study shows that the skills used most frequently by graduates were those of the EMT and the Occupational Health aspects of the course.
- Advanced skillsets were used rarely even when deployed overseas.
- In developing future CMT courses it is recommended that the EMT and Occupational health skill sets are emphasised, with streamlining of the advanced skillset elements to focus the skills are most relevant in an overseas environment.
- It would be advantageous to the Defence Forces for systems be put in place to recognise qualifications and support graduating students in practicing and using their new skills.

## Introduction

The Diploma in Military Medical Care (DMMC) has been developed and delivered by the UCD Centre for Emergency Medical Science (CEMS) in conjunction with the Irish Defence Forces (DF) and has been running from 2016 to 2019. The Irish Defence Forces have a long standing and well recognised role in UN mandated peacekeeping and humanitarian missions involving troop deployments to austere or high threat environments with specific medical support needs; the DF also sustains home based or ‘garrison’ roles which require quite different inputs from medical services. Recent overseas missions have included Army deployments to Syria, Mali and Lebanon and Naval Service deployments to the Mediterranean.

The mission of the DF Medical Corps is to ensure the coordination and provision of medical, dental and pharmaceutical support to the DF in the execution of its roles^1^. One of the key roles of the DF is “to participate in multinational peace support, crisis management and humanitarian relief operations in support of the United Nations and under UN mandate, including regional security missions authorised by the UN”^2^. At home and overseas, this means working in Primary Care or Role 1 medical units. Role 1 medical support is described by NATO as:

*“* … *providing first aid, immediate lifesaving measures, and triage. Additionally, it will contribute to the health and well-being of the unit through provision of guidance in the prevention of disease, non-battle injuries, and operational stress. Normally, routine sick call and the management of minor sick and injured personnel for immediate return to duty are a function of this level of care*.*”* ^3^

The DF identified the need for an innovative training programme to equip its non-medical staff to address these roles, in the context of a role known as ‘Combat Medical Technician’(CMT), leading to the development of the UCD DMMC programme.

The CMT/DMMC initiatives are among a range of significant recent developments within the DF Medical Corps. In October 2015 Military Medicine within the DF was recognised by the Irish Medical Council as a new medical specialty.^4^ It was recognised that this specialty was needed to provide “*a sustainable model for workforce planning in the Medical Corps by helping to address the medical requirements of the Defence Forces, and also recognised the unique skill sets required of Medical Officers in their work at home and overseas”*.^*5*^

All DMMC programme participants are members of the Irish Defence Forces. Training takes place at Health Sciences Belfield, the Curragh Camp and at clinical placements in DF Occupational Health units, within the UCD GP Tutor network, in National Ambulance Service placements and at Emergency Departments around the country. The intensive one-year programme equips graduates to undertake the demanding role of CMT at home and in deployed settings.

The DMMC programme includes four key components;

i. emergency Medical Technician (EMT) course and qualification,
ii. advanced pre-hospital skills course (for use in deployed settings under medical supervision),
iii. occupational health technical skills course,
iv. tactical military medical care skills course.

The ‘EMT skill set’ includes patient assessment, primary survey of ABC’s (Airway, Breathing, Circulation assessment), CPR/AED use, key medical, trauma and paediatric emergency care, tourniquet use and some medication administration. The ‘Advanced skill set’ (under medical supervision) includes IV/IO access and advanced life support in cardiac arrest, fluid resuscitation/tranexamic acid use, needle decompression, traction splinting and analgesia. The ‘Occupational health skill set’ includes blood pressure (BP) measurements, 12-lead ECG, phlebotomy, immunisation, spirometry, audiometry and medical record keeping. Tactical military medical skills were taught and assessed at the Curragh by DF instructors.

The EMT part of the course is run over six weeks at the beginning of the DMMC course. All students must pass their EMT exams and become registered with the Pre-Hospital Emergency Care Council (PHECC), to continue with the Diploma course. Since the first intake in Sept 2016, 55 Diploma graduates and 16 EMT-only graduates have been trained; of the 55 DMMC graduates 22 (40%) now work in the medical unit of the DF and the reminder in other units of the DF. Overall 25/55 (45%) are in non-medical Army units and two (4%) have left the DF. ^6^

For most of these students, this is their first experience of University learning. The Diploma is intended as the initial entry qualification for DF personnel wishing to join the Medical Corps.

This study looks at the opinions and experiences of the Defences Forces staff members who graduated from the DMMC and EMT courses at UCD from 2016-2019. It is hoped that this information can be used to improve the teaching and learning experience for future DF candidates at UCD.

## Methods

The Clinical Learning Environment and Supervision (CLES) Tool^7^ has been acknowledged as a valid and reliable instrument to evaluate clinical learning environments for nursing students. It has been adapted in several countries and contexts including for medical students in a primary healthcare setting^8^. In this context it was adapted to assess the learning environment for DF students at UCD and on clinical placements in ambulance services, GP surgeries, hospitals and military medical facilities. The CLES tool explored the areas of teaching, clinical placements and the overall learning experience (Figure 2). The respondents were asked to mark on a scale of one to five, whether they disagreed (one and two on the scale), were neutral (three on the scale) or agreed (four and five on the scale) with the statements. The mean value for each response is also shown.

**Figure 1.**
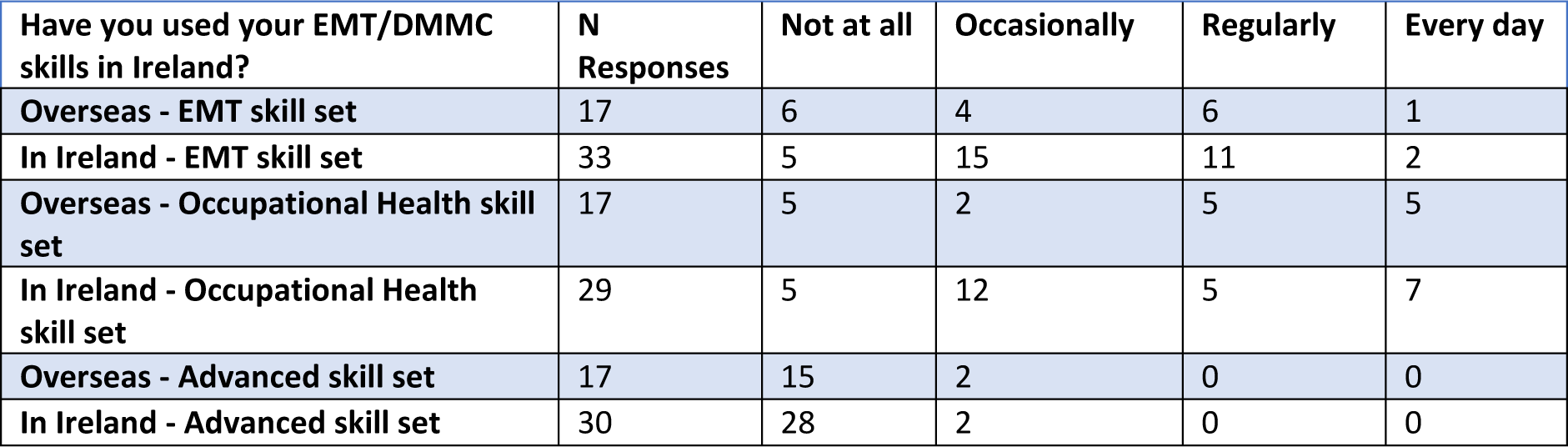
Use of Skill sets and Home and Overseas.

**Figure 2.**
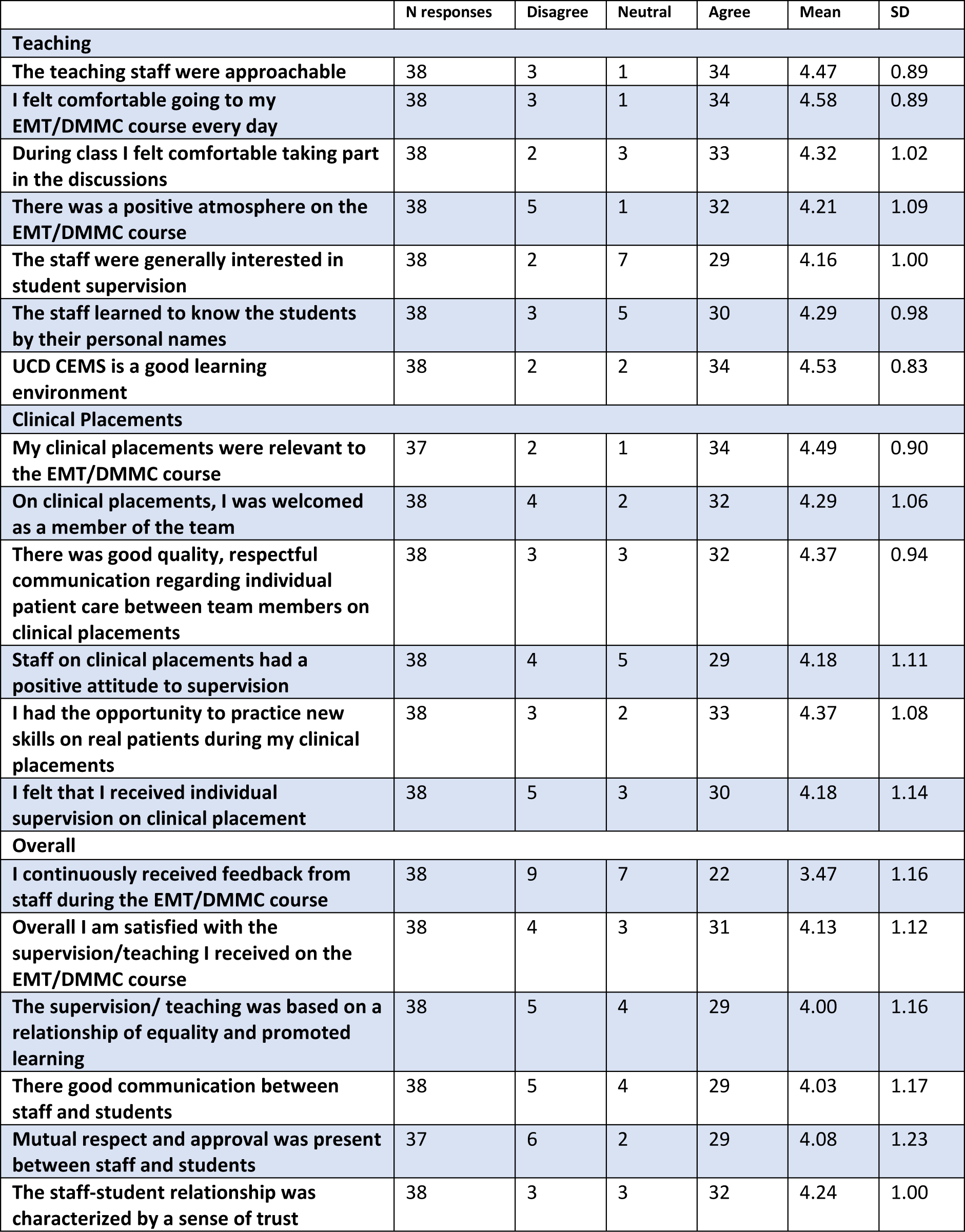
Educational CLES Rating of the course.

The adapted tool was incorporated into an anonymous online survey which was sent to the personal emails of 71 graduates from the DMMC and EMT-only courses from 2016 to 2019. Permission for the study was granted by the Defence Forces Medical School and UCD Human Research Ethics Committee.

## Results

There was a 53% response rate with 38 responses from 71 emails sent. The demographic profile of respondents showed that the majority, 34 (89%) were male, and 50% were aged between 25-34 years old. Regarding the use of new skills, 16 (42%) report using their new skills in their daily work. Nine respondents (24%) have been deployed overseas since the course; some had been overseas prior to taking part in the DMMC course. Of those deployed, all reported using at least some of their new medical skills while overseas.

Figure 1 outlines the use of EMT, advanced and occupational skill sets both in Ireland and overseas. The occupational health and EMT skill sets were used more often. The advanced skill sets were used only occasionally by a minority of graduates with most respondents reporting they had not used these advanced skills at all.

Student feedback using the adapted CLES tool was very positive in relation to the DMMC programme.

With regards to teaching, there was a high correlation between perceived staff interest in student supervision and a positive atmosphere on the course. The relevance of the clinical placements and opportunities to practice skills on real patients were also highly correlated. Overall, most statements were marked as four or five (agree) on the five-point scale. The only area identified by slightly lower mean score of 3.47 was in continuous staff feedback to students. However, this did not correlate with communication between staff and students, which was considered very good overall.

## Discussion

The results show that only 16 (42%) of respondents report using their DMMC/EMT skills in their daily work. Some Diploma graduates were not required to make a career move to a medical unit following the course, as the course was open to all Defence Forces personnel, including more senior non-commissioned officers from other units. Regardless, all graduates faced the challenge that much of their new skill set was not immediately applicable on a day-to-day basis within the DF for use in an Irish healthcare context. This results in loss of skills, less than ideal allocation of resources, and frustration for the new graduates in not being able to maintain or improve upon their new skillset.

The study shows that the skills used most frequently were those of the EMT and the Occupational Health aspects of the course. Advanced skillsets were used rarely even when deployed overseas. The feedback regarding the course was very positive overall regarding teaching and clinical placements. However, there was room for improvement in the area of continuous staff feedback to students.

It is important to bear in mind that the graduate’s perception of the course is heavily influenced by their clinical placement experience. A placement with opportunities to practice key skills and work as part of an inclusive team, with a chance of returning to that team after the course, would more likely provide a positive experience. Conversely, clinical placements that did not readily provide these opportunities, for example, where phlebotomy or other skills were the domain of other professions (as is the case in some of the military facilities), and where skills could not be practiced, were more likely to provide a negative experience.

Strengths of the study include the CLES tool, which is validated for use in many clinical learning environments including primary care. This study also provides both quantitative and qualitative feedback on the course.

A weakness of the study was that 33 (47%) of graduates did not respond. Their opinions and use of skills may be entirely different from those that responded. Only 9 out of 38 respondents reported deployment overseas since EMT/DMMC course, however there were 17 responses regarding use of skills overseas. This suggests that students may have been deployed overseas before the course. The survey could have been worded differently to differentiate between these responses.

It is hoped that the information gained from this study will be useful in designing future courses in collaboration with the Irish Defence Forces. It is recommended that the EMT and Occupational health skill sets are emphasised, with streamlining of the advanced skillset elements to focus on IV/IO access, definitive airway management and analgesia, as these advanced skills are most relevant in an overseas environment. There could be additional opportunities for staff-student feedback incorporated as part of the formative assessment of the course. Most importantly, it would be advantageous to the Defence Force for systems be put in place to recognise qualifications and allow graduating students to practice and use their new skills.

## Data Availability

Data are available upon reasonable request to

